# Meningococcal vaccination in adolescents and adults induces bactericidal activity against hyperinvasive complement-resistant meningococcal isolates

**DOI:** 10.1101/2022.05.20.22275303

**Authors:** Milou Ohm, Janine J. Wolf, Debbie M. van Rooijen, Linda J. Visser, Willem R. Miellet, Rob Mariman, Krzysztof Trzciński, Anne-Marie Buisman, Fiona R.M. van der Klis, Guy A.M. Berbers, Mirjam J. Knol, Nina M. van Sorge, Gerco den Hartog

**Author notes:** **Corresponding author:** dr. Gerco den Hartog, Centre for Infectious Disease Control Netherlands, National Institute for Public Health and the Environment (RIVM), Netherlands. **Alternate corresponding author:** Milou Ohm, Centre for Infectious Disease Control Netherlands, National Institute for Public Health and the Environment (RIVM), Netherlands.

## Abstract

**Background:** Complement-mediated killing is critical in the defense against meningococci. During a recent outbreak of invasive meningococcal serogroup W disease (IMD-W) in the Netherlands, the predominant isolates belonged to clonal complex (cc) 11, which may suggest a role for cc11-assocated traits in complement resistance. We investigated complement resistance of invasive and carriage meningococcal isolates of different serogroups and lineages. In addition, we investigated whether vaccine-induced antibodies can overcome resistance to complement-mediated killing.

**Methods:** We analyzed IMD isolates (n=56) and carriage isolates (n=19) of different serogroups and clonal lineages in the serum bactericidal antibody (SBA) assay using pooled serum from unvaccinated and vaccinated individuals. Furthermore, we determined meningococcal serogroup W geometric mean titers (GMTs) and protection levels with the routinely-used non-cc11 isolate and hyperinvasive cc11 isolates using individual serum samples from adolescents and adults 5 years postvaccination.

**Results:** The hyperinvasive IMD isolates showed high variation in their resistance to complement-mediated killing when pooled serum from unvaccinated individuals was used (median 96, range 2-1,536). When pooled sera from vaccinated individuals was used, all isolates were killed. The minimum spanning tree revealed moderate clustering of serogroup and cc, while complement resistance did not. While a significantly lower GMT was observed against cc11 meningococcal serogroup W (MenW) compared to a non-cc11 MenW isolate in vaccinated adults but not in adolescents, we found no differences in the proportion protected between these isolates.

**Conclusions:** These data show that vaccine-induced antibodies are effectively inducing complement-mediated killing of complement-resistant hyperinvasive and carriage meningococcal isolates.

**Short summary:** Meningococcal isolates of hyperinvasive lineages are resistant to complement-mediated killing but vaccine-induced antibodies effectively kill these invasive isolates.

## INTRODUCTION

The Gram-negative diplococcus *Neisseria meningitidis* - also known as meningococcus - is a human-specific bacterium that colonizes the upper respiratory tract (1). In rare cases, it behaves as an opportunistic pathogen (2) causing invasive meningococcal disease (IMD) often presented as meningitis or sepsis (3). The reason why one person is an asymptomatic carrier while another becomes severely ill remains largely unknown. IMD mainly affects otherwise healthy individuals, although complement deficiencies and splenic dysfunction/asplenia are known to predispose to IMD. In addition, individuals with these predispositions often show a reduced immune response to vaccination (4, 5).

IMD has been endemic with several major outbreaks across the world (6). The most recent outbreak of IMD in Europe was caused by serogroup W meningococci. Highest incidence rates were observed in the United Kingdom and the Netherlands, but an increased number of IMD serogroup W (IMD-W) cases was observed in most European countries. In the Netherlands, IMD-W incidence rates up to 1.2 per 100,000 were reported in 2018 – in contrast to rates below 0.05 per 100,000 before the outbreak in 2010-2014 – and an unusually high case fatality rate of 26% was observed in 14-24 year-olds (7).

Characterization of IMD-W outbreak isolates showed a dominance of meningococci belonging to the hyperinvasive clonal complex (cc) 11 (8). This lineage was also implicated in an outbreak among Hajj pilgrims that started in 2000 and Hajj-associated outbreaks in the meningitis belt thereafter (9). Moreover, an IMD-W outbreak in South America that emerged in 2003 and the IMD-C outbreak in Europe in early ‘00 were both dominated by meningococci belonging to cc11 (10, 11). However, other lineages such as cc23 and cc41/44 also contribute to epidemics and hyperendemic situations (12).

The first line of defense against meningococci is provided by the mucosal barrier. In the rare event of the bacterium passing the mucosal barrier, the complement system is an important component of the innate immune system that is able to quickly eliminate invading bacteria (13). The complement system plays a major role in preventing IMD, exemplified by the fact that deficiencies in components of its terminal pathway are associated with recurrent *N. meningitidis* infection. Complement can be activated by bacterial surfaces, however, many bacterial pathogens including *N. meningitidis* have acquired mechanisms to evade complement-mediated killing. For example, binding of factor H by the factor H binding protein (fHbp) inhibits activation of the alternative complement pathway through C3 (14). Serogroup-specific antibodies can induce complement-mediated killing of meningococci and overcome these pathogenic immune-evasion mechanisms (15). Antibodies against surface proteins including porins may be acquired naturally following colonization and may add to bactericidal activity of serum (16).

Although the importance of specific lineages is recognized in epidemics, little is known on how it relates to resistance to complement-mediated killing. We aimed to assess whether IMD isolates with distinct polysaccharide capsules and belonging to a variety of ccs differ in their intrinsic resistance to complement-mediated killing using sera from unvaccinated individuals. Furthermore, we investigated to what extent antibodies induced by a MenACWY-TT vaccine are able to overcome potential complement resistance of hyperinvasive meningococcal isolates.

## METHODS

### Isolate collection

Invasive meningococcal isolates (n=56) were selected to represent diverse clinical backgrounds, serogroups and lineages (Table 1). IMD is a notifiable disease in the Netherlands and invasive isolates are typed by the National Reference Laboratory for Bacterial Meningitis (NRLBM), Amsterdam, the Netherlands. Isolates selected for this study were collected between 2001 and 2018 and were serogrouped by Ouchterlony gel diffusion (17). Clinical background information (Supplementary Table 1) was collected through the national surveillance system for IMD in the Netherlands (18). Since MenA is not prevalent in the Netherlands, the selection was restricted to serogroup C, W and Y. In addition, a diverse selection of meningococcal serogroup CWY and nongroupable carriage isolates (serogrouped with Meningococcus Agglutination Sera [Oxoid©](19)) of varying cc (n=19) were included from a carriage study among students performed in the Netherlands in 2018 (20) (Table 1).

**Table 1.** Characteristics of selected isolates.

| Isolation | Serogroup | Clonal complex | total (n) | Age, median (range) | Clinical manifestation |  |  |  |  |
| --- | --- | --- | --- | --- | --- | --- | --- | --- | --- |
|  |  |  |  |  | pneumonia | sepsis | meningitis | arthritis | unspecified |
| Disease<br>n=56 | C | 8 | 2 | 2.5 (2-3) | 0 | 0 | 2 | 0 | 1 |
|  |  | 11 | 11 | 19 (0-72) | 0 | 3 | 7 | 1 | 4 |
|  |  | other | 4 | 50 (1-53) | 0 | 2 | 2 | 0 | 0 |
|  |  | total | 17 | 19 (0-72) | 0 (0%) | 5 (29%) | 11 (65%) | 1 (6%) | 5 (29%) |
|  | W | 11 | 18 | 43 (0-73) | 3 | 7 | 4 | 2 | 3 |
|  |  | 22 | 6 | 35 (1-79) | 0 | 4 | 4 | 0 | 1 |
|  |  | other | 2 | 63 (56-70) | 0 | 0 | 0 | 1 | 1 |
|  |  | unknown | 2 | 34 (0-68) | 0 | 2 | 0 | 0 | 0 |
|  |  | total | 28 | 43 (0-79) | 3 (11%) | 13 (46%) | 8 (29%) | 2 (7%) | 5 (18%) |
|  | Y | 23 | 6 | 68 (18-78) | 1 | 2 | 2 | 1 | 0 |
|  |  | 167 | 2 | 45 (19-71) | 0 | 0 | 2 | 0 | 0 |
|  |  | unknown | 3 | 66 (16-70) | 1 | 1 | 0 | 0 | 1 |
|  |  | total | 11 | 67 (16-78) | 2 (18%) | 3 (27%) | 4 (36%) | 1 (9%) | 1 (9%) |
| Carriage<br>n=19 | C | total | 1 | N/A | no symptoms |  |  |  |  |
|  | W | total | 4 | N/A | no symptoms |  |  |  |  |
|  | Y | total | 10 | N/A | no symptoms |  |  |  |  |
|  | NG | total | 4 | N/A | no symptoms |  |  |  |  |

NG = nongroupable; N/A = not applicable. Note that one patient may present with more than one clinical manifestation.

### Serum bactericidal antibody assay

The serum bactericidal antibody (SBA) assay was performed using pooled serum as well as individual sera and baby rabbit complement as complement source (21, 22). The serum pools consisted of sera from 1) unvaccinated adults and 2) adolescents and young adults five years after a single dose of the MenACWY-TT vaccine (Supplementary Table 2) (23, 24). We determined the resistance to killing of invasive and carriage isolates in the absence (serum pool 1) and presence (serum pool 2) of vaccine-induced antibodies. A detailed description of the assay can be found in Supplementary Information. Briefly, all tested isolates were grown on gonococcal (GC)-agar plates at 37°C and 5% CO_2_ and harvested at exponential growth phase. The SBA titer was determined by plating serial two-fold dilutions of serum on Colombia blood agar plates. When the SBA titer of the serum pool fell below 4, i.e. the isolate was not killed in a serum dilution of 1:2, we considered the isolate to be highly resistant to complement-mediated killing. If the titer exceeded the limit of quantitation (titer ≥262,144), i.e. the bacterium lacked resistance to complement-mediated killing entirely, we assigned a titer of 262,144. The SBA titer of the serum pool was assessed with the average of two independent experiments performed in duplicate. When the two independent measurements differed by more than two dilutions, additional replicates were performed in order to obtain a reproducible titer.

The individual sera from adolescents (n=54) and middle-aged adults (n=50) were collected five years after a single dose of MenACWY-TT in two clinical studies (NL4518 and NL7735) (23). The sera were tested with the default MenW non-cc11 target isolate (cc22, strain MP01240070) and the default MenW cc11 target isolate (2160326) used in our laboratory for SBA assays. Additionally, we tested a randomly selected subset (n=22) of sera from the adolescent cohort to determine the SBA titer against three additional serogroup W cc11 isolates to explore the variation in resistance to killing among different isolates belonging to this specific cc (isolate #30, isolate #35, isolate #49; Supplementary Table 1 and 3). The internationally accepted correlate of protection (SBA titer ≥8) was used for analysis of the individual sera, with the bactericidal titer defined as the dilution of sera that yielded ≥50% killing after 60 minutes incubation (23-25). A value of 2 was assigned when the titer fell below the cut-off of the assay (titer <4).

### Phylogenetic analyses

We used core genome multilocus sequence typing (cgMLST) to construct a minimum spanning tree in order to explore possible phylogenetic relationships for invasiveness of various genetic lineages. Whole genome sequencing (WGS) was performed for all *N. meningitidis* isolates included in this study. Genomic DNA was extracted using Genelute Bacterial Genomic DNA kit (NA2120, Sigma). Libraries were prepared using Illumina DNA prep kit (20018705, Illumina) and paired-end sequenced (2×150 bp) on an Illumina NextSeq platform (Illumina), following the manufacturer’s protocols. Read quality analysis and *de novo* assembly was performed with the Juno-assembly v2.0.2 pipeline (25). Briefly, fastq quality assessment and filtering were performed using FastQC and FastP. Genomes were assembled using SPAdes and curated with QUAST, CheckM and Bbtools. Subsequently, isolates were typed with MLST v2.19.0 (26) as well as chewBBACA (v2.8.5) (27) for the 1605 gene loci included in the Neisseria cgMLST scheme from pubMLST (28). cgMLST data were used to construct a minimum spanning tree with GrapeTree (v2.1) (29). *In silico* capsule typing was performed as described by Marjuki et al. (30).

### Statistical analyses

Statistical analyses were performed using GraphPad Prism v9 and SPSS Statistics v28. Differences in log-transformed SBA titers between serogroups for the different serum pools were tested with a mixed-effect analysis for paired data with Sidak’s correction for multiple comparisons. Differences in log-transformed SBA titers between the different clinical manifestations were determined with a Kruskal-Wallis test with Dunn’s multiple comparisons correction. The interaction between serogroup and cc with log-transformed SBA titer as dependent variable was investigated with a generalized linear model (GLM). We determined the geometric mean titers (GMTs) per age group of the individual sera for serogroup-specific SBA titers. Differences between age groups in log-transformed SBA titers of the individual sera were determined with a Wilcoxon matched-pairs signed rank test for paired data using Pratt’s method to account for tied values within a sample. Differences in proportions of protected individuals following vaccination were determined by a Chi-square or Fisher’s exact test as appropriate. Differences between the GMT of a subset of individual sera for different MenW cc11 isolates were compared with a Friedman’s ANOVA test with Dunn’s multiple comparisons correction. A p-value of <0.05 was considered statistically significant.

## RESULTS

### Description of selected isolates

Characteristics for all tested meningococcal isolates with details on the serogroup and source of isolation including clinical background of patients for all invasive isolates are summarized in Table 1 (Supplementary Table 1: data per isolate). For the collection of invasive isolates, the median age of patients was 45 years (range 0-79). Meningitis was the most commonly reported clinical manifestation (23/56, 41%). Invasive isolates causing clinical manifestations such as pneumonia and arthritis were also included in the selection. Half of the invasive isolates belonged to serogroup W (28/56, 50%) of which 18 (64%) and 6 (21%) belonging to cc11 and cc22 respectively. The majority of MenC isolates belonged to cc11 (10/17, 59%), while there was no MenY cc11 available for inclusion. Among MenY isolates, cc23 was the primary cc (6/11, 55%), followed by cc167 (2/11, 18%). Nongroupable isolates were identified among the carriage isolates.

### Bacterial killing by pooled serum from unvaccinated and vaccinated individuals

We found a significant difference (p<0.0001) in SBA titers against invasive isolates between the serum pools of unvaccinated and vaccinated individuals (Figure 1A). In the presence of vaccine-induced antibodies, all isolates (invasive and carriage) were killed. All carriage isolates except for one were killed by complement when exposed to the pool of serum from unvaccinated individuals. For 7 out of 19 (37%) carriage isolates a titer above the limit of quantitation (≥262144) was determined (Figure 1A). All isolates survived in presence of high serum concentrations when active complement was absent. Seventeen out of 56 (30%) invasive isolates were highly resistant to complement-mediated killing and survived in the presence of high serum concentrations from unvaccinated individuals. The majority of resistant isolates belonged to serogroup W (13/17, 76%) which also showed the lowest SBA GMTs, followed by MenY (Figure 1B). In contrast to MenW and MenY isolates, all MenC isolates showed limited resistance against the pool of serum from unvaccinated individuals. A significant difference was found between SBA titers determined with serum from unvaccinated individuals for carriage isolates compared with invasive isolates that caused sepsis (p=0.02) but not for isolates that caused meningitis, pneumonia or arthritis or that resulted in death (Figure 1C). If the non-resistant nongroupable isolates were excluded from the analyses, the significant difference for the clinical manifestation sepsis disappeared. No trend regarding the age of the patient was observed among resistant isolates. To identify a possible relation between complement resistance and ccs, data were plotted according to the isolate’s cc (Figure 2). All ccs showed wide variation in SBA titers in absence of vaccine-induced antibodies (Figure 2A), which reduced after vaccination with no evidence for difference between ccs after vaccination (Figure 2B). GLM revealed that serogroup (p<0.001) and cc (p=0.011) of invasive isolates associated with SBA titers in absence but not in presence of vaccine-induced antibodies.

**Figure 1.**
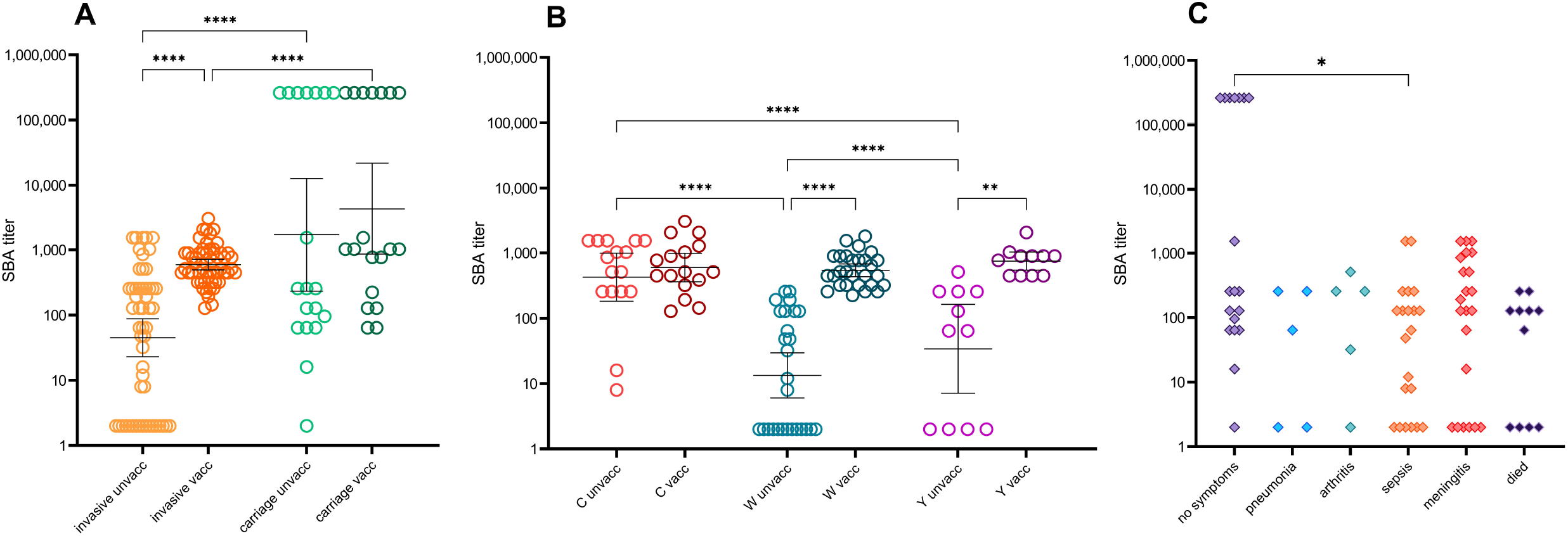
Serum bactericidal antibody titers of invasive and carriage meningococcal isolates determined with pooled serum from unvaccinated individuals and pooled serum from vaccinated individuals. A) all invasive and all carriage isolates for both serum pools, B) invasive isolates per serogroup C, W and Y for both serum pools, C) SBA titers per clinical manifestation for the pool of serum from unvaccinated individuals. Statistical analyses performed with a mixed-effect analysis with Sidak’s correction for multiple comparison on log-transformed data. Geometric mean SBA titers with 95% confidence intervals are shown in panel A and B for each group. SBA = serum bactericidal antibody; unvacc = tested with pool of serum from unvaccinated individuals, vacc = tested with pool of serum from vaccinated individuals. *p<0.05; ** p<0.01; *** p<0.001; **** p<0.0001.

**Figure 2.**
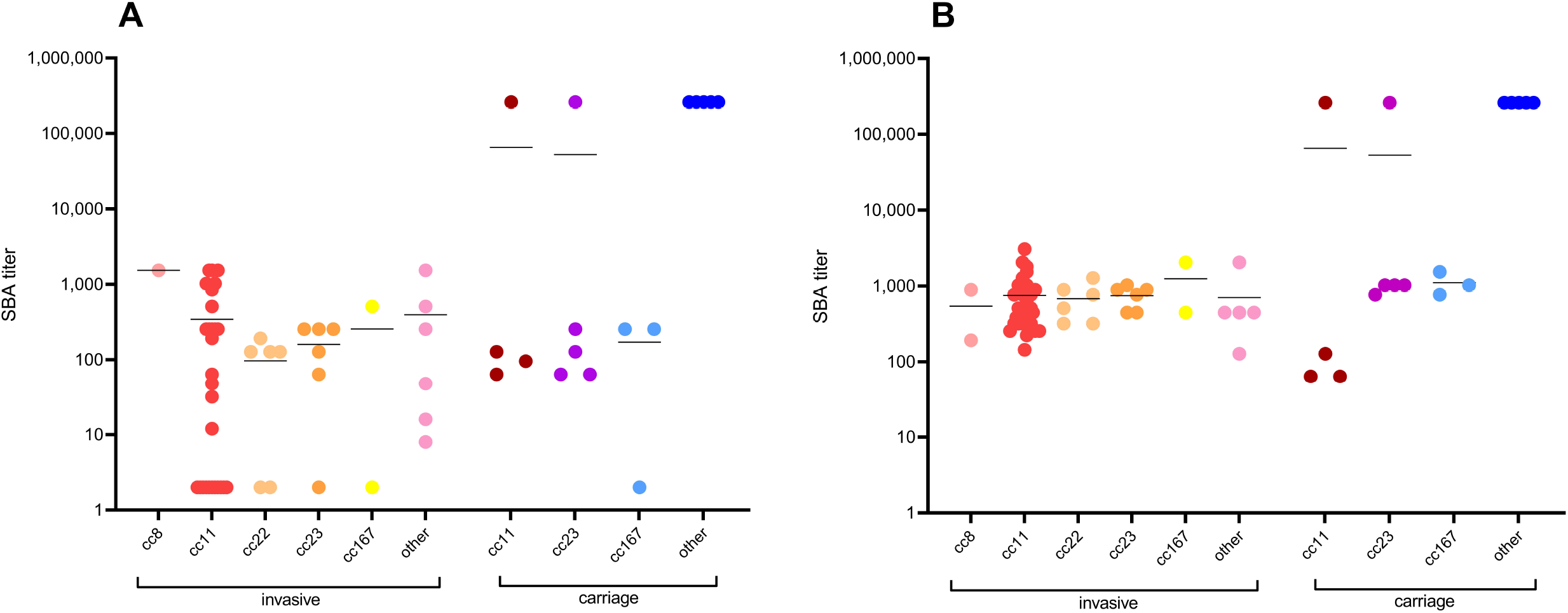
Serum bactericidal antibody titers of invasive and carriage meningococcal isolates per clonal complex determined with A) pooled serum from unvaccinated individuals and B) pooled serum from vaccinated individuals. SBA = serum bactericidal antibody; cc = clonal complex. Geometric mean SBA titers are shown for each clonal complex. For three isolates (n=1 for cc8 and n=2 for other), a result was available for only one of the serum pools.

### Phylogenetics

In order to explain the observed differences in complement resistance, we performed WGS and cgMLST of the isolates. No apparent clustering was observed for the source of isolation (Figure 3A). Despite heterogeneity within serogroups and ccs, several serogroups and cc subclusters in close genetic distance were observed (Figure 3B). Although overall classes of SBA did not cluster (Figure 3C), the cgMLST analyses revealed clustering of invasive MenW cc11 as well as cc22 isolates with medium to high resistance to killing, while invasive MenC cc11 isolates clustered with low to no resistance to killing. Clustering for non-cc11 isolates (including MenY isolates) was not revealed other than the already established separation based on serogroup.

**Figure 3.**
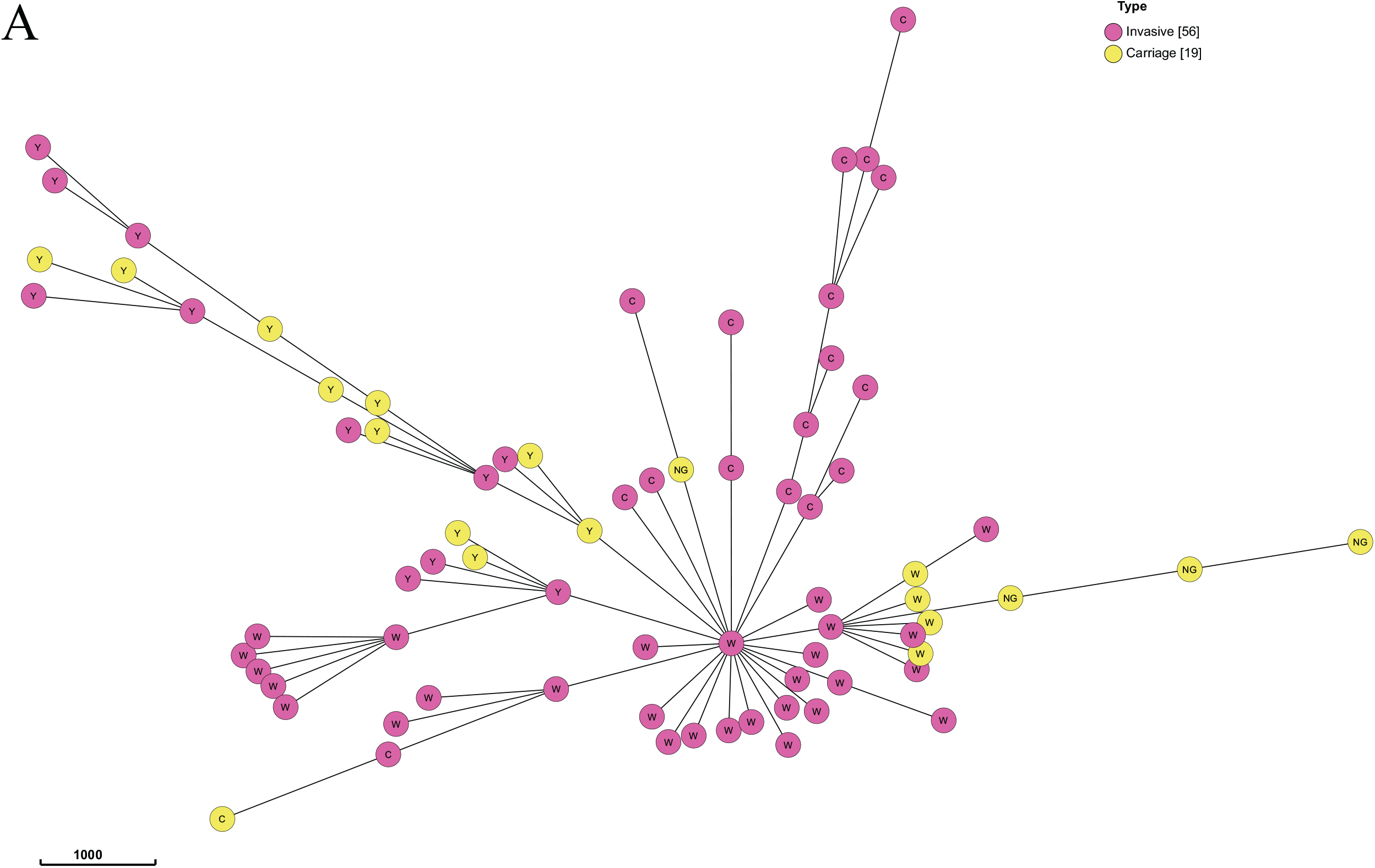
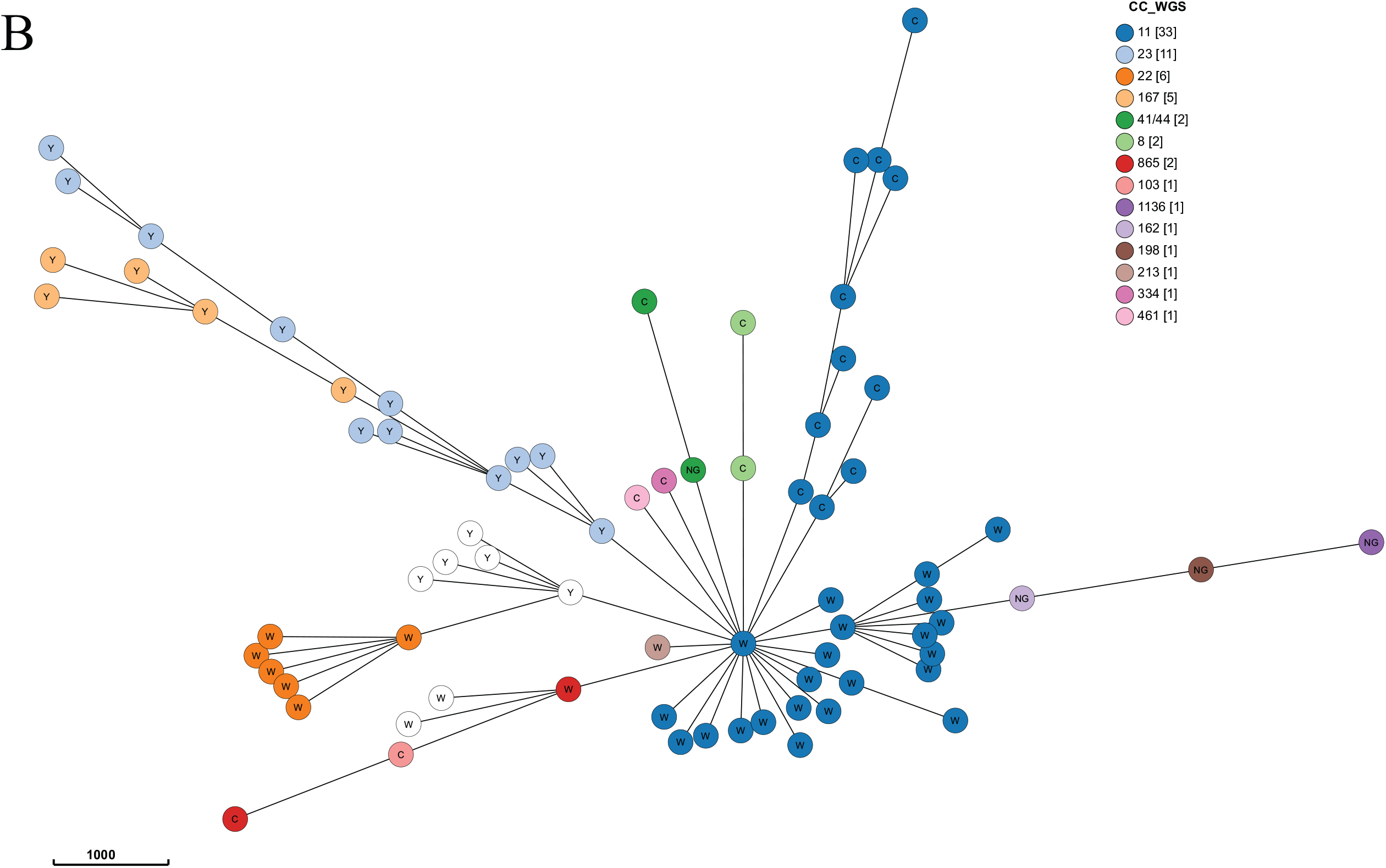
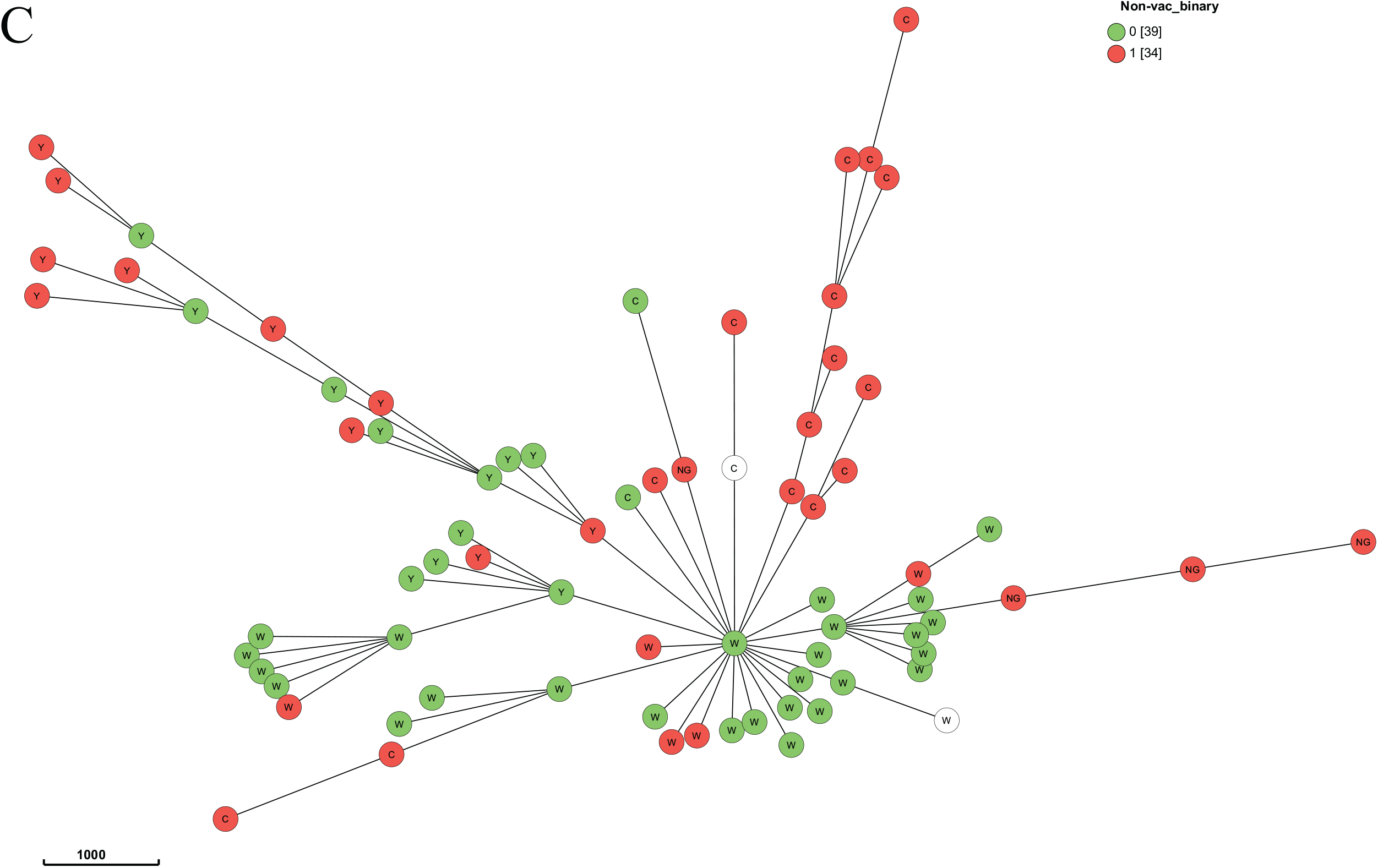
Minimum spanning trees of cgMLST data per A) source of isolation B) serogroup and clonal complex and C) serum bactericidal antibody (SBA) titer class* when tested with pooled serum of unvaccinated individuals. cc = clonal complex; WGS = whole genome sequencing; non-vac_binary = binary SBA titer class tested with pooled serum of unvaccinated individuals. *The SBA titer class as shown in C was assigned 0 (no to low resistance, in green) for a titer ≤128 and 1 (medium to high resistance, in red) for a titer >128. Every circle represents one isolate, the letter in the circle represents the serogroup. If a circle is white-colored, data is missing. There are five invasive isolates and two carriage isolates for which the ST does not belong to a cc, thus these values are missing. The scale of the branch length is shown on scale depicted in the left lower corner.

### Serum bactericidal titers in vaccinated individuals

To test the degree of individual protection (SBA titer ≥8) to cc11 MenW following vaccination, a MenW cc11 isolate and a MenW non-cc11 isolate were exposed to individual serum samples of adolescents and adults five years after MenACWY vaccination. A slightly higher GMT (p=0.03) was detected against the MenW non-cc11 isolate compared to the MenW cc11 isolate in adults but not in adolescents (Figure 4). However, the proportion of adults protected against both MenW isolates was similar (71% versus 66% with SBA titer ≥8 for the non-cc11 isolate and cc11 isolate, respectively). In a randomly selected subset of adolescents (n=22), we found either similar (isolate #35 and #49) or higher (isolate #30, p=0.004) GMTs to additionally tested MenW cc11 isolates in comparison to the non-cc11 isolate (Supplementary Table 3).

**Figure 4.**
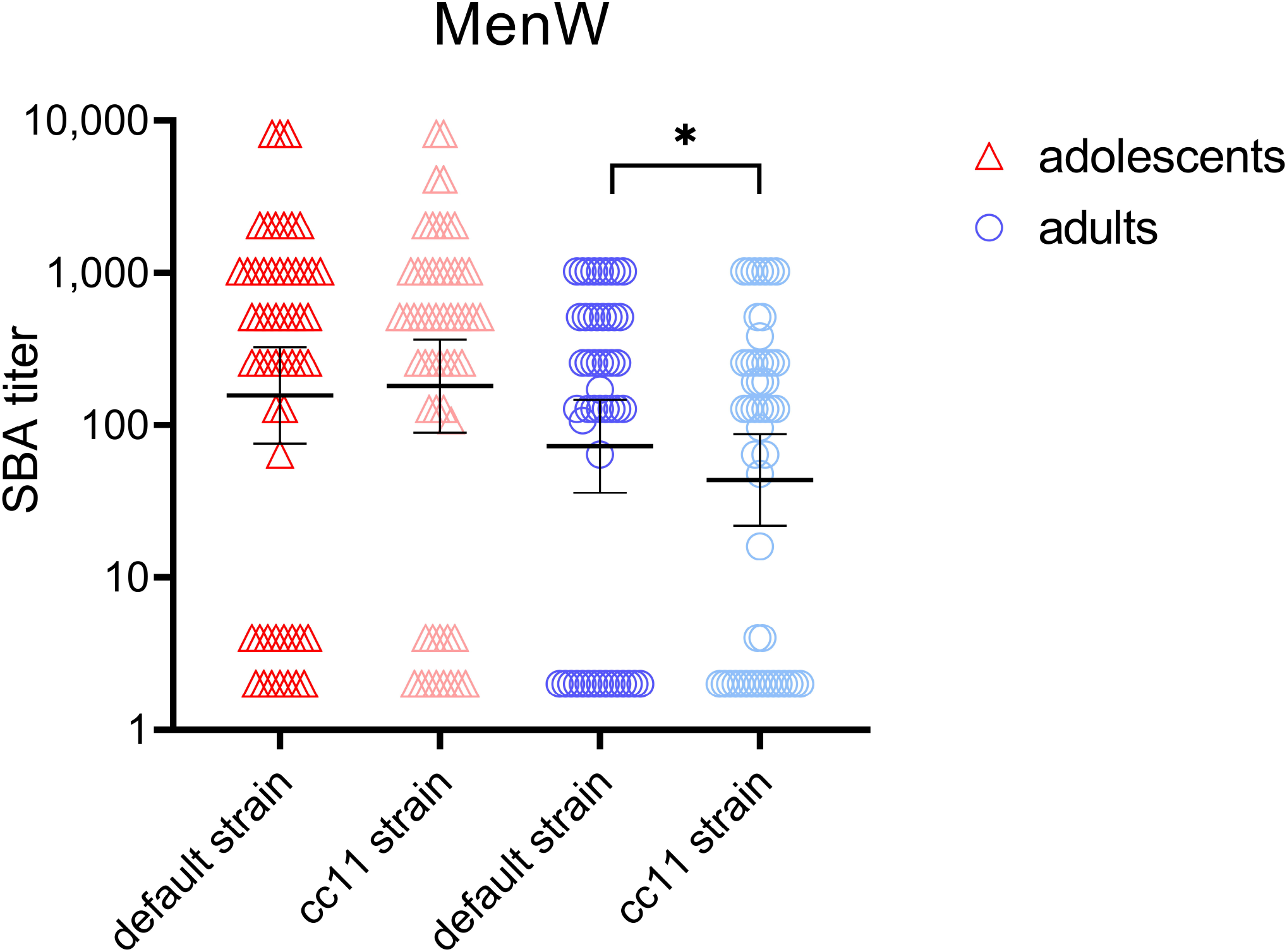
Serum bactericidal antibody titers with geometric mean titers in individual serum samples from vaccinated adolescents (n=54, red triangle) and adults (n=50 blue circle) against a serogroup W laboratory non-cc11 isolate and a serogroup W hyperinvasive cc11 isolate. *p<0.05 as determined with a Wilcoxon matched-pairs signed rank test using Pratt’s method. Bold lines are geometric mean SBA titers with 95% confidence intervals.

## DISCUSSION

In this study, we determined the resistance to complement-mediated killing of hyperinvasive clinical meningococcal isolates, and how this may be explained by the clonal complex. Our data showed that clinical isolates more often show resistance to complement-mediated killing compared to carriage isolates. Vaccine-induced antibodies overcome the resistance to killing of hyperinvasive meningococcal isolates. Hyperinvasive lineages with a specific cc were found to be in close genetic relationship. Five years postvaccination, vaccine-induced antibodies resulted in slight reduced SBA titers to MenW cc11 but not in a difference in the proportion of individuals with a SBA titer above the protective cut-off of 8.

In the presence of vaccine-induced antibodies, all tested isolates belonging to hyperinvasive lineages were killed. Since vaccination targets the polysaccharide capsule, which is largely independent of the clonal complex-associated genes, we did not expect a link between virulence and ccs when vaccine-induced antibodies were present. Epidemiological data previously suggested vaccine effectiveness against invasive isolates (31), of which we now provide mechanistic proof. While some ccs are implicated in IMD, others are rarely invasive and there is large heterogeneity between isolates belonging to the same cc. Within the invasive ccs, classes of SBA titers did not cluster. Yet, MenW cc11 isolates showed high resistance to killing while MenC cc11 isolates did not. The difference between serogroups might be explained by the colonization history of donors, with naturally-acquired antibodies in the serum from unvaccinated individuals (due to historical circulation of MenC in the Netherlands (32)) inducing bactericidal activity against MenC. Possibly, these antibodies – which might be targeted against the capsule but also other surface antigens – overcome the resistance to killing. Furthermore, it has previously been described that capsular polysaccharides are heterogeneous in their activation of the alternative pathway of complement, which may contribute to differences between serogroups in pathogenesis (33). Since isolates implicated in IMD also circulate through the population (34), it is not surprising that we also found one carriage isolate (belonging to cc167) to be resistant to complement-mediated killing with pooled serum from unvaccinated individuals.

Invasiveness of meningococci is thought to be determined – at least partly – by a combination of genetic characteristics of both the bacterium and the host (35, 36). Some ccs are known to be overrepresented in disease isolates, yet the STs itself are not necessarily involved in virulence. Virulence was previously found to be associated with the combination of cell surface antigens and the polysaccharide capsule, which is an essential antigen shielding the bacterium from host immunity but alone insufficient to confer full virulence (37). Evasion strategies involve several virulence factors such as the polysaccharide capsule, as well as fHbp, PorA, PorB and NspA next to some other molecules and factors involved in biofilm formation (13). In this study, although capsule expression of our isolates was confirmed, altered capsule expression through phase variation affecting complement sensitivity cannot be excluded. Furthermore, we used baby rabbit complement instead of human complement and therefore we could not investigate the four mentioned proteins – all related to factor H binding – above. Factor H binding protein – involved in regulation of the alternative pathway by inactivating C3 convertase - is species-specific and rabbit factor H does not bind to the surface of the meningococcal bacterium. Nevertheless, resistance varied among the different tested lineages which could be due to the combination of genes not reflected by the ST of each isolate (38) or other bacterial determinants such as genes involved in bacterial metabolism that could play a role in invasiveness (39). A genome-wide association analysis might reveal *N. meningitidis* pathogenicity genes, but our study was not powered for this type of analysis. Further research into the link between ccs and virulence factors and the association with bactericidal activity might elucidate how hyperinvasive meningococci evade the immune system.

We found a lower GMT in individual serum samples from adults (but not adolescents) to a cc11 isolate compared to an unrelated non-cc11 isolate (Figure 3). However, the proportion protected did not differ and most individuals unprotected (SBA titer <8) for the cc11 isolate were also found to be unprotected for the non-cc11 isolate. Additionally tested cc11 MenW isolates showed comparable titers in a subset of participants (Supplementary Table 3). The waning of long-term seroprotection after MenACWY vaccination is different between these age groups, as was previously studied in those individuals as part of a larger cohort (23).

This study has some limitations. Firstly, we used baby rabbit complement for the functional antibody assays and thus we might have yielded higher titers in the SBA assay than we would have with human complement. Furthermore, variation in the individual’s complement component - known as the complotype – alters complement activity of each individual thereby affecting disease risk (40). Secondly, we selected the isolates based on characteristics such as age, sex, clinical manifestations, and diversity of ccs. The results cover diversity in host-and genetic traits, but do not represent serogroup and lineage-specific epidemiology.

We conclude that vaccine-induced bactericidal antibodies allow bactericidal killing of a diverse set of clinically-relevant serogroup C, W and Y (hyper)invasive meningococci. Our results underline the importance of use of a meningococcal vaccination to provide protection against hyperinvasive lineages during an outbreak or hyperendemic situation.

## Data Availability

All data produced in the present study are available upon reasonable request to the authors

## Acknowledgements

We thank all involved lab technicians and Boas van der Putten from the NRLBM for their contribution to sample processing and analyses.

## Author contributions

M.O. and G.d.H. conceived and designed the study. M.J.K and N.M.v.S. were involved in data collection. G.A.M.B. and A.M.B. have supervised the longitudinal vaccination studies providing the serum samples. M.O., J.J.W. and D.M.v.R performed the serological analyses. L.V. and W.R.M., R.M. and K.T. contributed analytical tools and were involved in the genetic analyses. M.O., L.V. and G.d.H. interpreted and verified the data and made the figures. M.O. and G.d.H. and drafted the manuscript. All authors interpreted the data, critically reviewed the manuscript, and approved the final version.

## REFERENCES

1. Stephens DS. Biology and pathogenesis of the evolutionarily successful, obligate human bacterium Neisseria meningitidis. Vaccine. 2009;27:B71–B7.

2. Moxon ER, Jansen VAA. Phage variation: understanding the behaviour of an accidental pathogen. Trends in microbiology. 2005;13(12):563–5.

3. Caugant DA, Maiden MC. Meningococcal carriage and disease--population biology and evolution. Vaccine. 2009;27 Suppl 2:B64–70.

4. Balmer P, Falconer M, McDonald P, Andrews N, Fuller E, Riley C, et al. Immune response to meningococcal serogroup C conjugate vaccine in asplenic individuals. Infection and immunity. 2004;72(1):332–7.

5. Meerveld-Eggink A, de Weerdt O, de Voer RM, Berbers GA, van Velzen-Blad H, Vlaminckx BJ, et al. Impaired antibody response to conjugated meningococcal serogroup C vaccine in asplenic patients. European journal of clinical microbiology & infectious diseases : official publication of the European Society of Clinical Microbiology. 2011;30(5):611–8.

6. Pace D, Pollard AJ. Meningococcal disease: clinical presentation and sequelae. Vaccine. 2012;30 Suppl 2:B3–9.

7. Pluijmaekers A, de Melker H. The National Immunisation Programme in the Netherlands. Surveillance and developments in 2020-2021. Het Rijksvaccinatieprogramma in Nederland Surveillance en ontwikkelingen in 2020-2021: Rijksinstituut voor Volksgezondheid en Milieu RIVM; 2021.

8. Knol MJ, Hahné SJM, Lucidarme J, Campbell H, de Melker HE, Gray SJ, et al. Temporal associations between national outbreaks of meningococcal serogroup W and C disease in the Netherlands and England: an observational cohort study. Lancet Public Health. 2017;2(10):e473–e82.

9. Lucidarme J, Hill DM, Bratcher HB, Gray SJ, du Plessis M, Tsang RS, et al. Genomic resolution of an aggressive, widespread, diverse and expanding meningococcal serogroup B, C and W lineage. The Journal of infection. 2015;71(5):544–52.

10. Abad R, Lopez EL, Debbag R, Vazquez JA. Serogroup W meningococcal disease: global spread and current affect on the Southern Cone in Latin America. Epidemiology and infection. 2014;142(12):2461–70.

11. Safadi MA, Bettinger JA, Maturana GM, Enwere G, Borrow R, Global Meningococcal I. Evolving meningococcal immunization strategies. Expert review of vaccines. 2015;14(4):505–17.

12. Watkins ER, Maiden MC. Persistence of hyperinvasive meningococcal strain types during global spread as recorded in the PubMLST database. PloS one. 2012;7(9):e45349.

13. Lewis L, Ram S. Meningococcal disease and the complement system. Virulence. 2013;5.

14. Principato S, Pizza M, Rappuoli R. Meningococcal factor H binding protein as immune evasion factor and vaccine antigen. FEBS Letters. 2020;594(16):2657–69.

15. Frasch CE, Borrow R, Donnelly J. Bactericidal antibody is the immunologic surrogate of protection against meningococcal disease. Vaccine. 2009;27 Suppl 2:B112–6.

16. Pollard AJ, Frasch C. Development of natural immunity to Neisseria meningitidis. Vaccine. 2001;19(11):1327–46.

17. Ouchterlony O. Antigen-antibody reactions in gels. Acta Pathol Microbiol Scand. 1949;26(4):507–15.

18. Brandwagt DAH, van der Ende A, Ruijs WLM, de Melker HE, Knol MJ. Evaluation of the surveillance system for invasive meningococcal disease (IMD) in the Netherlands, 2004–2016. BMC infectious diseases. 2019;19(1):860.

19. Severin WPJ. Latex agglutination in the diagnosis of meningococcal meningitis. Journal of clinical pathology. 1972;25(12):1079–82.

20. Miellet WR, Mariman R, Pluister G, de Jong LJ, Grift I, Wijkstra S, et al. Detection of Neisseria meningitidis in saliva and oropharyngeal samples from college students. Sci Rep. 2021;11(1):23138.

21. Goldschneider I, Gotschlich EC, Artenstein MS. Human immunity to the meningococcus. II. Development of natural immunity. The Journal of experimental medicine. 1969;129(6):1327–48.

22. Maslanka SE, Gheesling LL, Libutti DE, Donaldson KB, Harakeh HS, Dykes JK, et al. Standardization and a multilaboratory comparison of Neisseria meningitidis serogroup A and C serum bactericidal assays. The Multilaboratory Study Group. Clinical and diagnostic laboratory immunology. 1997;4(2):156–67.

23. Ohm M, van Rooijen DM, Bonacic Marinovic AA, van Ravenhorst MB, van der Heiden M, Buisman AM, et al. Different Long-Term Duration of Seroprotection against Neisseria meningitidis in Adolescents and Middle-Aged Adults after a Single Meningococcal ACWY Conjugate Vaccination in The Netherlands. Vaccines (Basel). 2020;8(4).

24. van der Heiden M, Boots AMH, Bonacic Marinovic AA, de Rond LGH, van Maurik M, Tcherniaeva I, et al. Novel Intervention in the Aging Population: A Primary Meningococcal Vaccine Inducing Protective IgM Responses in Middle-Aged Adults. Frontiers in immunology. 2017;8(817).

25. Github Juno pipeline [Available from: https://github.com/RIVM-bioinformatics/Juno_pipeline.

26. Github MLST [Available from: https://github.com/tseemann/mlst.

27. Silva M, Machado MP, Silva DN, Rossi M, Moran-Gilad J, Santos S, et al. chewBBACA: A complete suite for gene-by-gene schema creation and strain identification. Microbial Genomics. 2018;4(3).

28. Bratcher HB, Corton C, Jolley KA, Parkhill J, Maiden MCJ. A gene-by-gene population genomics platform: de novo assembly, annotation and genealogical analysis of 108 representative Neisseria meningitidis genomes. BMC genomics. 2014;15(1):1138.

29. Zhou Z, Alikhan NF, Sergeant MJ, Luhmann N, Vaz C, Francisco AP, et al. GrapeTree: visualization of core genomic relationships among 100,000 bacterial pathogens. Genome Res. 2018;28(9):1395–404.

30. Marjuki H, Topaz N, Rodriguez-Rivera LD, Ramos E, Potts CC, Chen A, et al. Whole-Genome Sequencing for Characterization of Capsule Locus and Prediction of Serogroup of Invasive Meningococcal Isolates. Journal of clinical microbiology. 2019;57(3):e01609–18.

31. Ohm M, Hahné SJM, van der Ende A, Sanders EAM, Berbers GAM, Ruijs WLM, et al. Vaccine impact and effectiveness of meningococcal serogroup ACWY conjugate vaccine implementation in the Netherlands: a nationwide surveillance study. Clinical Infectious Diseases. 2021.

32. Bijlsma MW, Brouwer MC, Spanjaard L, van de Beek D, van der Ende A. A decade of herd protection after introduction of meningococcal serogroup C conjugate vaccination. Clinical infectious diseases : an official publication of the Infectious Diseases Society of America. 2014;59(9):1216–21.

33. Ram S, Lewis LA, Agarwal S. Meningococcal Group W-135 and Y Capsular Polysaccharides Paradoxically Enhance Activation of the Alternative Pathway of Complement*. Journal of Biological Chemistry. 2011;286(10):8297–307.

34. Ravenhorst MB, Bijlsma MW, van Houten MA, Struben VM, Anderson AS, Eiden J, et al. Meningococcal carriage in Dutch adolescents and young adults; A cross-sectional and longitudinal cohort study. Clinical microbiology and infection : the official publication of the European Society of Clinical Microbiology and Infectious Diseases. 2017.

35. Stabler RA, Marsden GL, Witney AA, Li Y, Bentley SD, Tang CM, et al. Identification of pathogen-specific genes through microarray analysis of pathogenic and commensal Neisseria species. Microbiology (Reading). 2005;151(Pt 9):2907–22.

36. Brouwer MC, de Gans J, Heckenberg SG, Zwinderman AH, van der Poll T, van de Beek D. Host genetic susceptibility to pneumococcal and meningococcal disease: a systematic review and meta-analysis. The Lancet Infectious diseases. 2009;9(1):31–44.

37. Joseph B, Schwarz RF, Linke B, Blom J, Becker A, Claus H, et al. Virulence evolution of the human pathogen Neisseria meningitidis by recombination in the core and accessory genome. PloS one. 2011;6(4):e18441.

38. Kugelberg E, Gollan B, Tang CM. Mechanisms in Neisseria meningitidis for resistance against complement-mediated killing. Vaccine. 2008;26 Suppl 8(6):I34–I9.

39. Watkins ER, Maiden MC. Metabolic shift in the emergence of hyperinvasive pandemic meningococcal lineages. Sci Rep. 2017;7:41126.

40. Harris CL, Heurich M, Cordoba SRd, Morgan BP. The complotype: dictating risk for inflammation and infection. Trends in Immunology. 2012;33(10):513-21.

