## Supplementary Material for "Meningococcal vaccination in adolescents and adults induces bactericidal activity against hyperinvasive complement-resistant meningococcal isolates"

**Supplementary Table 1. Characteristics, genetics and SBA titers for each meningococcal isolate.**

| **Isolate** | | | | **Clinical manifestation (0=no, 1=yes)** | | | | | | **MLST** | | **SBA titer** | |
| --- | --- | --- | --- | --- | --- | --- | --- | --- | --- | --- | --- | --- | --- |
| **Isolate** | **Type** | **Year of isolation** | **Serogroup*** | **Pneumonia** | **Bacteremia / sepsis** | **Meningitis** | **Arthritis** | **Other / Unspecified** | **Died** | **Clonal complex** | **ST** | **Serum pool unvaccinated** | **Serum pool vaccinated** |
| 1 | Invasive | 2018 | Y | 0 | 0 | 1 | 0 | 0 | 0 | 167 | 167 | 2 | 448 |
| 2 | Invasive | 2018 | Y | 0 | 0 | 0 | 0 | 1. Unspecified | 0 |  | 5436 | 2 | 896 |
| 3 | Invasive | 2018 | Y | 0 | 1 | 0 | 0 | 0 | 1 | 23 | 1655 | 128 | 448 |
| 4 | Invasive | 2018 | Y | 0 | 0 | 1 | 0 | 0 | 0 | 23 | 23 | 64 | 768 |
| 5 | Invasive | 2017 | Y | 0 | 0 | 1 | 0 | 0 | 0 | 167 | 167 | 512 | 2048 |
| 6 | Invasive | 2017 | Y | 1 | 0 | 0 | 0 | 0 | 0 |  | 5436 | 64 | 896 |
| 7 | Invasive | 2017 | Y | 0 | 1 | 0 | 0 | 0 | 0 |  | 5437 | 2 | 448 |
| 8 | Invasive | 2017 | Y | 1 | 0 | 0 | 0 | 0 | 0 | 23 | 23 | 256 | 1024 |
| 9 | Invasive | 2017 | Y | 0 | 1 | 0 | 0 | 0 | 0 | 23 | 23 | 256 | 896 |
| 10 | Invasive | 2017 | Y | 0 | 0 | 0 | 1 | 0 | 0 | 23 | 6800 | 256 | 896 |
| 11 | Invasive | 2017 | Y | 0 | 0 | 1 | 0 | 0 | 0 | 23 | 1655 | 2 | 448 |
| 12 | Invasive | 2001 | C | 0 | 0 | 1 | 0 | 0 | 0 | 11 | 11 | 1536 | 2048 |
| 13 | Invasive | 2001 | C | 0 | 0 | 1 | 0 | 1. Shock | 0 | 8 | 1372 | unknown | 896 |
| 14 | Invasive | 2001 | C | 0 | 1 | 0 | 0 | 1. Shock | 1 | 11 | 11 | 256 | 448 |
| 15 | Invasive | 2001 | C | 0 | 0 | 1 | 0 | 0 | 0 | 11 | 11 | 256 | 384 |
| 16 | Invasive | 2001 | C | 0 | 0 | 1 | 0 | 1. Shock | 1 | 11 | 2704 | 256 | 512 |
| 17 | Invasive | 2002 | C | 0 | 0 | 1 | 0 | 1. Shock | 0 | 11 | 11 | 1024 | 3072 |
| 18 | Invasive | 2017 | C | 0 | 0 | 1 | 0 | 0 | 0 | 11 | 11 | 853 | 512 |
| 19 | Invasive | 2017 | C | 0 | 1 | 0 | 0 | 0 | 0 | 11 | 16061 | 1536 | 768 |
| 20 | Invasive | 2017 | C | 0 | 1 | 0 | 0 | 0 | 0 | 41/44 | 14893 | 8 | 128 |
| 21 | Invasive | 2017 | C | 0 | 1 | 0 | 0 | 0 | 0 | 11 | 11 | 256 | 144 |
| 22 | Invasive | 2017 | C | 0 | 0 | 0 | 1 | 0 | 0 | 11 | 3035 | 512 | 320 |
| 23 | Invasive | 2018 | C | 0 | 1 | 0 | 0 | 0 | 0 | 103 | 5133 | 1536 | unknown |
| 24 | Invasive | 2018 | C | 0 | 0 | 1 | 0 | 0 | 0 | 11 | 11 | 1536 | 1280 |
| 25 | Invasive | 2001 | C | 0 | 0 | 1 | 0 | 0 | 0 | 334 | 2698 | 512 | 2048 |
| 26 | Invasive | 2001 | C | 0 | 0 | 1 | 0 | 0 | 0 | 8 | 2680 | 1536 | 192 |
| 27 | Invasive | 2001 | C | 0 | 0 | 1 | 0 | 1. Shock | 0 | 11 | 11 | 1024 | 1024 |
| 28 | Invasive | 2002 | C | 0 | 0 | 1 | 0 | 0 | 0 | 461 | 461 | 16 | 448 |
| 29 | Invasive | 2012 | W | 0 | 0 | 0 | 0 | 1. Unspecified | 0 | 22 | 1158 | 192 | 896 |
| 30 | Invasive | 2012 | W | 0 | 0 | 0 | 0 | 1. Unspecified | 0 | 11 | 11 | unknown | 1792 |
| 31 | Invasive | 2013 | W | 0 | 0 | 0 | 0 | 1. Unspecified | 0 | 11 | 11 | 2 | 448 |
| 32 | Invasive | 2013 | W | 0 | 0 | 0 | 0 | 1. Unspecified | 1 | 11 | 11 | 2 | 256 |
| 33 | Invasive | 2015 | W | 0 | 1 | 0 | 0 | 0 | 0 | 11 | 11 | 48 | 256 |
| 34 | Invasive | 2016 | W | 0 | 0 | 1 | 0 | 0 | 0 | 11 | 11 | 2 | 320 |
| 35 | Invasive | 2016 | W | 0 | 1 | 0 | 0 | 0 | 1 | 11 | 11 | 64 | 1536 |
| 36 | Invasive | 2016 | W | 1 | 0 | 0 | 0 | 0 | 0 | 11 | 11 | 2 | 768 |
| 37 | Invasive | 2016 | W | 0 | 1 | 0 | 0 | 0 | 1 | 11 | 11 | 2 | 768 |
| 38 | Invasive | 2016 | W | 1 | 0 | 0 | 0 | 0 | 0 | 11 | 11 | 2 | 320 |
| 39 | Invasive | 2017 | W | 0 | 0 | 1 | 0 | 0 | 1 | 11 | 11 | 2 | 256 |
| 40 | Invasive | 2017 | W | 0 | 0 | 1 | 0 | 0 | 0 | 11 | 11 | 192 | 320 |
| 41 | Invasive | 2017 | W | 0 | 1 | 1 | 0 | 0 | 1 | 22 | 3422 | 128 | 1280 |
| 42 | Invasive | 2017 | W | 1 | 0 | 0 | 0 | 0 | 0 | 11 | 11 | 256 | 1024 |
| 43 | Invasive | 2017 | W | 0 | 0 | 0 | 1 | 0 | 0 | 213 | 213 | 256 | 448 |
| 44 | Invasive | 2018 | W | 0 | 1 | 0 | 0 | 0 | 0 | 11 | 11 | 2 | 448 |
| 45 | Invasive | 2018 | W | 0 | 1 | 0 | 0 | 0 | 1 | 11 | 11 | 2 | 448 |
| 46 | Invasive | 2018 | W | 0 | 1 | 0 | 0 | 0 | 0 | 11 | 11 | 12 | 640 |
| 47 | Invasive | 2018 | W | 0 | 0 | 0 | 1 | 0 | 0 | 11 | 11 | 32 | 512 |
| 48 | Invasive | 2018 | W | 0 | 0 | 1 | 0 | 0 | 0 | 11 | 11 | 2 | 896 |
| 49 | Invasive | 2018 | W | 0 | 1 | 0 | 0 | 0 | 0 | 11 | 11 | 2 | 224 |
| 50 | Invasive | 2018 | W | 0 | 1 | 1 | 0 | 0 | 1 | 22 | 22 | 128 | 768 |
| 51 | Invasive | 2018 | W | 0 | 1 | 0 | 0 | 0 | 0 |  |  | 8 | 896 |
| 52 | Invasive | 2015 | W | 0 | 1 | 0 | 0 | 0 | 0 | 22 | 3422 | 2 | 512 |
| 53 | Invasive | 2016 | W | 0 | 0 | 1 | 0 | 0 | 0 | 22 | 1158 | 2 | 320 |
| 54 | Invasive | 2016 | W | 0 | 1 | 1 | 0 | 0 | 0 | 22 | 12236 | 128 | 320 |
| 55 | Invasive | 2017 | W | 0 | 0 | 0 | 0 | 1. Unspecified | 0 | 865 | 12256 | 48 | 448 |
| 56 | Invasive | 2018 | W | 0 | 1 | 0 | 0 | 0 | 1 |  | 9316 | 128 | 768 |
| 101 | Carriage |  | C (NG) | 0 | 0 | 0 | 0 | 0 | 0 | 865 | 3327 | 262144 | 262144 |
| 102 | Carriage |  | NG (C) | 0 | 0 | 0 | 0 | 0 | 0 | 41/44 | 1097 | 262144 | 262144 |
| 103 | Carriage |  | NG (NG) | 0 | 0 | 0 | 0 | 0 | 0 | 1136 | 1136 | 262144 | 262144 |
| 104 | Carriage |  | NG (NG) | 0 | 0 | 0 | 0 | 0 | 0 | 162 | 2153 | 262144 | 262144 |
| 105 | Carriage |  | NG (NG) | 0 | 0 | 0 | 0 | 0 | 0 | 198 | 823 | 262144 | 262144 |
| 106 | Carriage |  | W (NG) | 0 | 0 | 0 | 0 | 0 | 0 | 11 | 11 | 262144 | 262144 |
| 107 | Carriage |  | W (NG) | 0 | 0 | 0 | 0 | 0 | 0 | 11 | 11 | 128 | 64 |
| 108 | Carriage |  | W (NG) | 0 | 0 | 0 | 0 | 0 | 0 | 11 | 11 | 128 | 32 |
| 109 | Carriage |  | W (NG) | 0 | 0 | 0 | 0 | 0 | 0 | 11 | 11 | 96 | 64 |
| 110 | Carriage |  | Y (NG) | 0 | 0 | 0 | 0 | 0 | 0 | 167 | 767 | 2 | 768 |
| 111 | Carriage |  | Y (NG) | 0 | 0 | 0 | 0 | 0 | 0 | 23 | 1655 | 256 | 1024 |
| 112 | Carriage |  | Y (Y) | 0 | 0 | 0 | 0 | 0 | 0 |  | 5436 | 16 | 224 |
| 113 | Carriage |  | Y (Y) | 0 | 0 | 0 | 0 | 0 | 0 | 167 | 16465 | 768 | 1536 |
| 114 | Carriage |  | Y (NG) | 0 | 0 | 0 | 0 | 0 | 0 | 23 | 1655 | 128 | 768 |
| 115 | Carriage |  | Y (NG) | 0 | 0 | 0 | 0 | 0 | 0 | 23 | 10098 | 384 | 1024 |
| 116 | Carriage |  | Y (NG) | 0 | 0 | 0 | 0 | 0 | 0 |  | 16468 | 1536 | 128 |
| 117 | Carriage |  | Y (Y) | 0 | 0 | 0 | 0 | 0 | 0 | 167 | 167 | 256 | 1024 |
| 118 | Carriage |  | Y (NG) | 0 | 0 | 0 | 0 | 0 | 0 | 23 | 1655 | 4 | 1024 |
| 119 | Carriage |  | Y (NG) | 0 | 0 | 0 | 0 | 0 | 0 | 23 | 23 | 262144 | 262144 |

ST = sequence type; cnl = capsule null locus. *Serogroup for invasive isolates was determined with Ouchterlony gel diffusion, for carriage isolates with serolatex agglutination assay. The letter between the brackets after each serogroup of all carriage isolates represents the genogroup as predicted by WGS. Note: if a field is blank, the ST of the isolate does not belong to a cc. All isolates were collected in the Netherlands.

**Supplementary Table 2. Details on the two pools of serum with information on donors and concentration of polysaccharide-specific IgG concentration*.**

|  |  | Serum pool 1 | Serum pool 2 |
| --- | --- | --- | --- |
| vaccination status of donors |  | Unvaccinated | Vaccinated 5 years prior to serum collection |
| n of donors |  | 10 | 11 |
| age of donors |  | 50-65 years | 15-20 years (10-15 at time of vaccination) |
| sex of donors (% female) |  | 80% (8 out of 10) | 45% (5 out of 11) |
| Weighted mean concentration of polysaccharide-specific IgG* (µg/mL) | MenC | 0.44 | 16.29 |
|  | MenW | 0.33 | 10.17 |
|  | MenY | 0.22 | 22.83 |

*determined with the multiplex immunoassay as described by De Voer et al. (1).

1. de Voer RM, van der Klis FR, Engels CW, Rijkers GT, Sanders EA, Berbers GA. Development of a fluorescent-bead-based multiplex immunoassay to determine immunoglobulin G subclass responses to Neisseria meningitidis serogroup A and C polysaccharides. Clinical and vaccine immunology : CVI. 2008;15(8):1188-93.

**Supplementary Table 3. Serum bactericidal antibody titers for one MenW non-cc11 isolate and four MenW cc11 isolates in a subset (n=22) of individual serum samples from adolescents five years after MenACWY vaccination.**

| **Sample no** | **non-cc11 (strain MP01240070)** | **Isolate 2160326** | **Isolate #30** | **Isolate #35** | **Isolate #49** |
| --- | --- | --- | --- | --- | --- |
| 1 | 256 | 256 | 512 | 256 | 256 |
| 2 | 1024 | 2048 | 2048 | 1024 | 1024 |
| 3 | 1024 | 1024 | 1024 | 1024 | 1024 |
| 4 | 512 | 512 | 1024 | 256 | 256 |
| 5 | 2 | 2 | 128 | <16 | <16 |
| 6 | 1024 | 1024 | 2048 | 512 | 1024 |
| 7 | 128 | 128 | 128 | <16 | <16 |
| 8 | 2048 | 2048 | 4096 | 1024 | 1024 |
| 9 | 4 | 2 | <16 | <16 | <16 |
| 10 | 1024 | 1024 | 1024 | 1024 | 1024 |
| 11 | 512 | 512 | 512 | 512 | 256 |
| 12 | 2 | 2 | <16 | <16 | <16 |
| 13 | 256 | 512 | 512 | 256 | 256 |
| 14 | 1024 | 1024 | 2048 | 2048 | 1024 |
| 15 | 1024 | 1024 | 2048 | 2048 | 512 |
| 16 | 256 | 512 | 512 | 128 | 256 |
| 17 | 2048 | 2048 | 4096 | 2048 | 1024 |
| 18 | 512 | 512 | 2048 | 512 | 512 |
| 19 | 1024 | 1024 | 2048 | 1024 | 512 |
| 20 | 512 | 512 | 512 | 512 | 512 |
| 21 | 256 | 512 | 1024 | 1024 | 256 |
| 22 | 256 | 256 | 256 | 256 | 128 |
| GMT* with 95% CI | 281 (113-700) | 309 (119-807) | 563 (246-1289) | 256 (99-660) | 205 (85-498) |

GMT = geometric mean titer; CI = confidence interval. Detailed information on the isolate can be found in Supplementary Table 1. *When a value of <16 was determined, a value of 4 was assigned for statistical analyses.

**Supplementary Information**

**Serum bactericidal antibody (SBA) assay: method**

The target isolate (stock culture stored at -80°C in Greaves freezing solution containing bovine serum albumin faction V, monosodium glutamate, glycerol and sterile water) was streaked with an inoculation loop on a gonococcal (GC)-agar plate (enriched with IsoVitaleX, BD Biosciences©) and incubated overnight at 37°C and 5% CO2 to obtain isolated colonies. The isolate was subcultured by spreading 5-8 colonies with a sterile swab over the entire surface of another GC-plate followed by an incubation of 4 hours at 37°C and 5% CO2. The cells were suspended in 5mL of D-PBS (Dulbecco’s PBS, ThermoFisher©, containing 0.50 mM MgCl2, 0.90 mM CaCl2, 0.33 mM pyruvate and 5.55 mM D-glucose) and set at absorbance of 0.300 at 600nm with a spectrophotometer. The bacterial suspension [dilution 1] was diluted in D-PBS in three steps (100 µl of dilution 1 in 1900 µL D-PBS [dilution 2], 50 µL of dilution 2 into 1950 µL D-PBS [dilution 3] and finally 300 µL of dilution 3 in 5700 µL D-PBS) to yield ~40-50 CFU per 10 µL. The baby rabbit complement (stored at -80°C) was freshly thawed and added to the bacterial suspension to reach a final concentration of complement of 20%. The mixture was added to the serially two-fold prediluted serum (heat-inactivated for 30 minutes at 56°C, starting dilution of 4) in D-PBS in a 96-wells plate. Each microtiter plate contained two control wells, per isolate when tested with pooled serum or per individual serum sample: one well with buffer, bacteria and active complement without serum, and one well with buffer, bacteria and heat-inactivated test serum without active complement. The microtiter plate was incubated for 60 minutes at 37°C without extra CO2. The average number of CFU added to each well before incubating (T0) was determined by plating 6 lanes of 10 µl of the bacteria/complement mixture onto a Colombia blood agar plate (Oxoid©) and allowing the mixture to flow down the plate (tilt method) with a multichannel pipette. After incubation of the microtiter plate, samples were plated as described for T0. The SBA titer of each serum sample (either pooled serum or an individual serum sample) was expressed as the serum dilution that yielded 50% killing compared to the CFU present at T0 (before incubation with serum) (1).

1.	Maslanka SE, Gheesling LL, Libutti DE, Donaldson KB, Harakeh HS, Dykes JK, et al. Standardization and a multilaboratory comparison of Neisseria meningitidis serogroup A and C serum bactericidal assays. The Multilaboratory Study Group. Clinical and diagnostic laboratory immunology. 1997;4(2):156-67.
